# The Utility of Optical Coherence Tomography Angiography Biomarkers in Detecting Diabetic Retinopathy

**DOI:** 10.64898/2026.04.22.26351527

**Authors:** Kieran Kumanan, Abolfazl Hassani, Muhammad Husnain, Elias Papaefstratiou, Jose J. Estevez

**Affiliations:** Caring Futures Institute, College of Nursing and Health Sciences, Flinders University, Bedford Park, Adelaide, South Australia, Australia

**Keywords:** Diabetic Retinopathy, Angiography, Associations, Risk Factors, Maculopathy, Diagnostic Accuracy

## Abstract

**Purpose:** To evaluate associations between optical coherence tomography angiography (OCT-A) metrics and diabetic retinopathy (DR) and compare their discrimination against conventional clinical risk factors.

**Methods:** In this cross-sectional study, 108 adult eyes (right eye if both eligible) with diabetes were recruited from tertiary ophthalmology/optometry clinics. DR was clinically graded using ETDRS categories and dichotomised as no DR vs ≥ mild NPDR (primary outcome). Macular 6×6 mm OCT-A (Zeiss AngioPlex) was acquired; scans with signal strength >7 and without major artefact were included. Quantitative metrics from the superficial capillary plexus included vessel density (VD) and perfusion density (PD) (central/inner/outer/full regions); structural OCT measures and FAZ parameters were secondary. Associations with ≥ mild NPDR were assessed using multivariable logistic regression adjusted for age, sex, HbA1c, and diabetes duration. Discrimination was evaluated with ROC curves/AUC (95% CI) and DeLong comparisons of AUCs.

**Results:** DR was present in 63% of eyes. DR was associated with lower VD (central, inner, outer, full) and lower PD (central, inner, full) (all p≤0.04). After adjustment, central VD (OR 0.82, 95% CI 0.68–0.98) and central PD (OR 0.92, 95% CI 0.86–0.99) remained independently associated with DR. The OCT-A model outperformed the clinical model (AUC 0.73 vs 0.60); the combined model yielded AUC 0.76.

**Conclusion:** VD and PD from the superficial plexus are independently associated with DR and show superior discrimination versus conventional clinical factors alone, supporting OCT-A as an adjunct for earlier DR detection.

## Introduction

Diabetic retinopathy (DR) is the most common microvascular complication of diabetes and a leading cause of preventable vision loss.^1^ Globally, an estimated 159 million individuals are affected by some form of DR, including approximately 28 million with vision-threatening endophenotypes such as proliferative DR and diabetic macular oedema (DMO).^2^

In DR, chronic hyperglycaemia induces both microangiopathy and neurodegeneration, leading to impaired autoregulation, reduced capillary perfusion, and retinal ischemia. Early pathological changes including pericyte loss, capillary basement membrane thickening, and ganglion cell loss precede and drive the development of clinically visible lesions.^3^ Early detection of DR is critical for enabling timely, targeted interventions. For instance, early detection of DR may prompt optimisation of modifiable risk factors, such as glycaemic control and blood pressure, recognised to reduce the likelihood of DR development and progression.^3^

Despite the need for early detection, current DR grading systems utilising conventional imaging modalities, such as funduscopic examination, often miss subtle microvascular changes. In contrast, optical coherence tomography angiography (OCT-A), has the capacity to detect subtle changes. OCT-A is a non-invasive imaging technique that provides high-resolution visualisation of the retinal vascular architecture. It detects motion contrast by comparing sequential B-scans of the same retinal location, attributing differences to erythrocyte movement within vessels.^4^ Unlike two-dimensional fundus fluorescein angiography, OCT-A is not obscured by hyper-fluorescence from dye leakage, enabling acquisition and quantitative analysis of parameters such as volumetric data, foveal avascular zone (FAZ) dimensions, perfusion metrics and microvascular alterations in both the superficial capillary plexus (SCP; comprising vasculature within the retinal nerve fibre and ganglion cell layer) and deep capillary plexuses (DCP).^5^

There is growing interest in OCT-A for detecting subtle microvascular changes that precede clinically apparent DR; however, whether OCT-A metrics alone reliably indicate DR or outperform systemic risk factors requires further investigation. Therefore, this study aims to investigate the associations between OCT-A metrics and the presence of DR and compare their discriminative performance against conventional systemic risk factors.

## Materials and Methods

### Study design and participants

The current study is a cross-sectional observational study. Participant recruitment occurred between February 2024 and July 2025. Participants who met the clinical selection criteria were recruited from Flinders ophthalmology and optometry diabetes eye clinics. Inclusion criteria comprised of confirmed diagnosis of adults with type 1 or 2 diabetes with or without DR, and the acquisition of 6 × 6 mm OCT-A images centred on the fovea with a signal strength index >7. Exclusion criteria included the presence of severe media opacity; previous macula or retinal diseases; glaucoma; other ocular diseases causing visual impairment; or any retinal treatment within six months prior to imaging, including intraocular surgery, anti-VEGF injections, ocular steroid administration, or retinal photocoagulation. Additional exclusion criteria were severe segmentation errors, motion artefacts, false flow signals, uncorrected astigmatism, defocus, or incorrect OCT instrument positioning. If both eyes met the inclusion criteria, the right eye was selected for OCT-A imaging to ensure consistency and avoid selection bias. OCT-A analyses were conducted using one imaged eye per participant, while DR severity was defined from the worst eye across both eyes to reflect the systemic nature of diabetes and to minimise bias from between-eye correlation. Ethical approval was obtained from the Southern Adelaide Local Health Network Human Research Ethics Committee (#HRE00228). All participants received written and verbal information about the study and provided written consent. The study adhered with the National Statement on Ethical Conduct in Human Research and the Declaration of Helsinki.

### Protocol and procedures

Research coordinators systematically collected data via questionnaires and medical records on clinical variables including age, sex, socioeconomic factors, diabetes duration and type. The most recent haemoglobin A1c (HbA1c) measurement was extracted from pathology records. Participants underwent macular imaging using the Zeiss Cirrus 5000 HD-OCT with AngioPlex ™ OCT-A system (Carl Zeiss Meditec, Dublin, CA, USA) after dilation with Tropicamide 1% and Phenylephrine 2.5%. DR clinical signs were graded by the examining clinician, following clinical interpretation of the 17 levels of the Early Treatment Diabetic Retinopathy Study (ETDRS) severity scale^6^ into no retinopathy, minimal non-proliferative DR (NPDR), mild NPDR, moderate NPDR, severe NPDR, and proliferative DR. Clinically significant macular oedema (CSMO), defined by the ETDRS^6^ as retinal thickening within 500 µm of the foveal centre, hard exudates within 500 µm with adjacent thickening, or thickening ≥1 disc area within one disc diameter of the fovea, were excluded to minimise imaging artefacts and preserve OCT-A reliability. However, DMO, defined by the ETDRS^6^ as retinal thickening in the macular area not meeting the CSMO criteria was included.

### OCT and OCT-A Metrics

The Zeiss Cirrus HD-OCT model 5000 with AngioPlex™ has a speed of 68,000 A-scans per second and central wavelength of 840LJnm.^7^ Structural OCT imaging was performed using the Macular Cube 512 × 128 scan protocol to acquire cross-sectional images of the retina. Quantitative metrics extracted from these scans included central foveal thickness (CFT), average macular thickness, and macular cube volume (mm³). OCT-A imaging was conducted using 6 × 6 mm macular scans, which were automatically segmented to the SCP and divided into nine subfields according to the ETDRS; the DCP was not measured.^8^ Blood flow was detected using the optical microangiography-complex algorithm (**Figure 1**).^9^ From these flow maps, the following quantitative OCT-A metrics were derived: perfusion density (PD), defined as the percentage (%) of the region of interest occupied by flow-positive pixels; vessel density (VD), defined as the total length of perfused vessels per mm², expressed in mm^-1^; FAZ area, defined as the area (mm²) of the largest non-perfused central region; FAZ perimeter, defined as the length (mm) of the FAZ boundary; and FAZ acircularity index, defined as the degree of deviation from a perfect circle. Motion artifacts are also minimised using the FastTrac™ retinal-tracking technology.^7,9^

**FIGURE 1:**
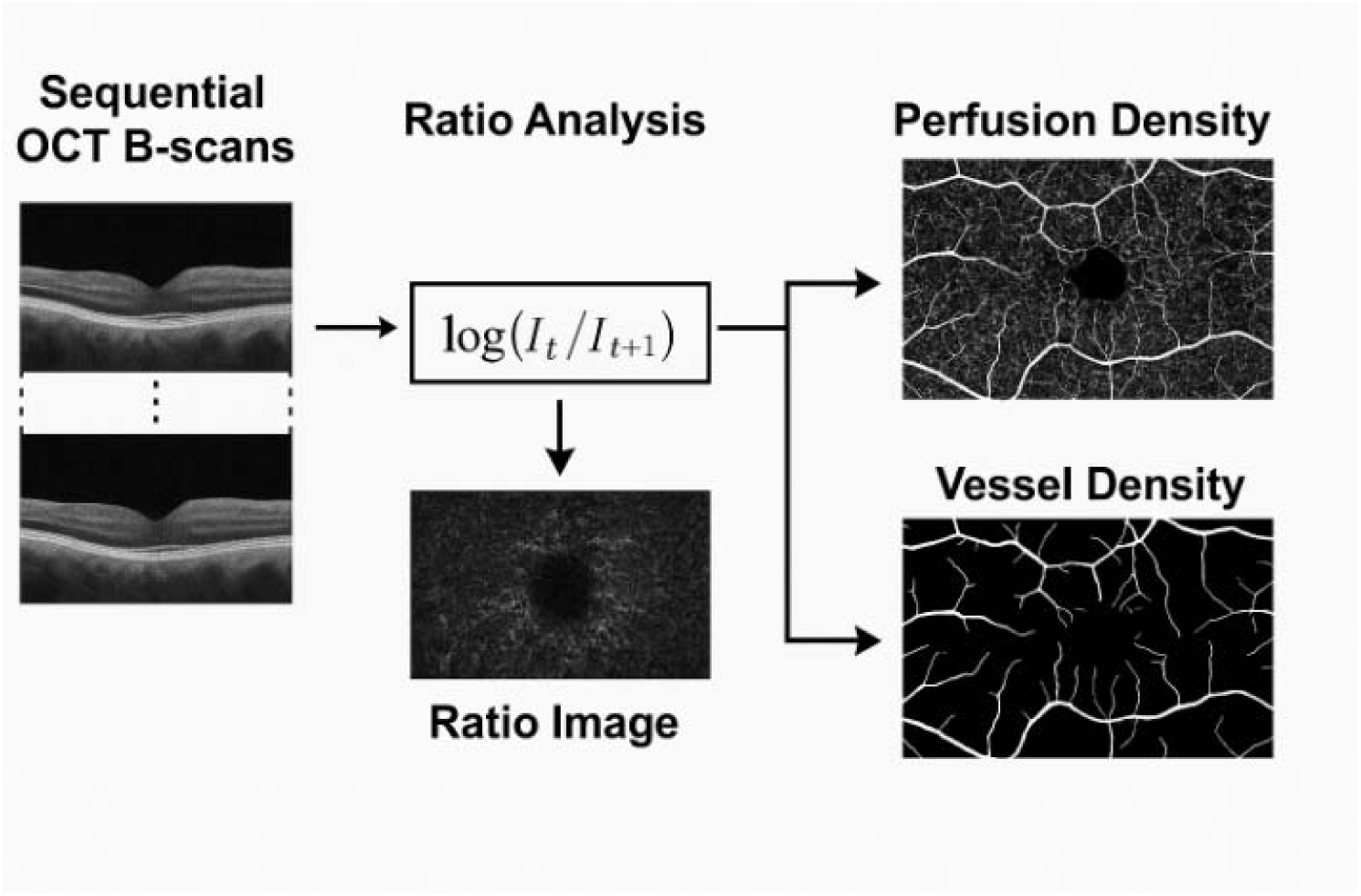
Log-intensity motion contrast between sequential B-scans calculated using the formula: R(x,y) = log[I□(x,y) / I□□□(x,y)]. Static tissue is characterised by similar signal intensities in sequential scans (I□ ≈ I□□□), producing a log ratio close to zero and no flow signal, whereas moving red blood cells produced differing signal intensities (I□ ≠ I□□□), resulting in a log ratio not equal to zero and a positive flow signal. Pixels with high temporal variance in this log-ratio signal were classified as perfused vessels, generating the OCT-A flow map (9).

### Statistical Analysis

Analyses were conducted at the participant level. Right and left eye grades were harmonised, and an overall DR severity was defined as the worse eye. DMO was derived analogously. Primary outcome was ≥ mild NPDR (binary); the full ordinal DR severity distribution was summarised descriptively. OCT-A availability was defined by a valid scan; OCT-A metrics included FAZ, PD and VD across central/inner/outer/full regions, and structural OCT measures. Visual acuity was analysed as logMAR. The FAZ area was rescaled per 0.1 mm² for interpretability. Continuous data are reported as mean ± SD and compared with two-sample t-tests (Wilcoxon when indicated), categorical data with Pearson’s χ² (Fisher’s exact when sparse). Associations with ≥ mild NPDR were examined using logistic regression. A clinical model (age, sex, HbA1c, diabetes duration), an OCT-A model (candidate OCT-A metrics within the OCT-A subgroup, with collinearity checks), and a combined model (clinical + OCT-A) were constructed. Collinearity was evaluated using variance inflation factors (VIF > 5) and correlation matrices, and pre-specified central and inner macular regions as primary predictors to limit multiplicity and reduce overfitting. Results are presented as odds ratios (ORs) with 95% confidence intervals (CIs). Discrimination was evaluated using ROC curves and AUCs (95% CIs), with paired AUC comparisons by DeLong’s test. A sensitivity analysis was performed restricting to participants where the imaged eye was the worse (or equal) eye. Analyses were hypothesis-driven, focusing on pre-specific macular subfields (central and inner) shown in prior studies to be most sensitive to early DR. Accordingly, no correlation for multiple comparisons was applied to these primary analyses; secondary regional comparisons were considered exploratory. Two-sided p < 0.05 was considered statistically significant. Analyses were performed in Stata (v18, StataCorp).

## Results

### Patient characteristics

A total of 108 right eyes from 108 participants with diabetes were included in the OCT-A analysis. The mean (SD) age was 58.3 (16.8) years, with a mean diabetes duration of 21.1 (12.2) years and a mean HbA1c of 7.6% (1.4). 40 eyes (37%) had no DR, and 68 eyes (63%) had ≥ mild NPDR. Among those with ≥ mild DR, 48% were classified as mild NPDR, 34% as moderate NPDR, 4% as severe NPDR, and 10 (14.7%) eyes had co-existing DMO. Most participants had type 2 diabetes (72%). Demographic and clinical characteristics are summarised in **Table 1**.

**TABLE 1.** Cohort characteristics overall and by diabetic retinopathy status.

| Cohort characteristics | Total Cohort<br>(n=108) | No DR<br>(n=40) | ≥ Mild NPDR(n=68) | P value* |
| --- | --- | --- | --- | --- |
| **Demographic and clinical factors** |  |  |  |  |
| Age (years), mean (SD) | 58.3 (16.8) | 56.7 (19.7) | 59.1 (15.6) | 0.48 |
| Sex, Male, n (%) | 72 (66.7) | 22 (55.0) | 50 (73.5) |  |
| Sex, Female, n (%) | 36 (33.3) | 18 (45.0) | 18 (26.5) |  |
| Diabetes duration (years), mean (SD) | 21.1 (12.2) | 20.2 (14.9) | 22 (10.2) | 0.46 |
| HbA1c (%), mean (SD) | 7.60 (1.4) | 7.4 (0.8) | 7.8 (1.3) | 0.26 |
| **Diabetes diagnosis, mean (SD)** |  |  |  |  |
| Diabetes type, n (%) |  |  |  | 0.65 |
| Type 1 | 29 (26.9) | 12 (30) | 17 (25) |  |
| Type 2 | 78 (72.2) | 28 (70) | 50 (73.5) |  |
| Other | 1 (0.9) | 0 (0) | 1 (1.5) |  |
| **Socioeconomic Factors** |  |  |  |  |
| Education (HS or less), n (%) | 39 (36.1) | 12 (30) | 27 (39.7) |  |
| Employed, n (%) | 43 (39.8) | 16 (40) | 27 (39.7) |  |
| Weekly income (< \$500), n (%) | 8 (7.4) | 5 (12.5) | 3 (4.4) | |
| Annual income (< \$37,000), n (%) | 17 (15.7) | 5 (12.5) | 12 (17.6) | |
| **Disease severity** |  |  |  |  |
| Overall DR (worse eye), n (%) |  |  |  | <0.001 |
| No DR | 35 (32.4) | 35 (87.5) | 0 (0) |  |
| Minimal | 5 (4.6) | 5 (12.5) | 0 (0) |  |
| Mild | 38 (35.2) | 0 (0) | 38 (55.9) |  |
| Moderate | 26 (24.1) | 0 (0) | 26 (38.2) |  |
| Severe NPDR | 3 (2.8) | 0 (0) | 3 (4.4) |  |
| PDR | 1 (0.9) | 0 (0) | 1 (1.5) |  |
| Overall DMO (worse eye), n (%) |  |  |  | <0.001 |
| None | 98 (90.7) | 40 (100.0) | 58 (85.3) |  |
| Macular oedema | 10 (9.3) | 0 (0) | 10 (14.7) |  |
| *Abbreviations:*<br>DR, diabetic retinopathy; NPDR, non-proliferative diabetic retinopathy; PDR, proliferative diabetic retinopathy; DMO, diabetic macular oedema; HbA1c, glycated haemoglobin; SD, standard deviation; HS, high school; OD, oculus dexter (right eye); OS, oculus sinister (left eye).<br>*P-values are student's t-test or chi-squared test, where appropriate. |  |  |  |  |

### OCT-A parameter findings

Eyes with ≥ mild NPDR demonstrated significantly lower VD in the central (p<0.01), inner (p<0.01), outer (p = 0.04), and full macular regions (p = 0.02) compared with eyes without DR (**Table 2**). PD was also reduced in the central (p<0.01), inner (p = 0.01), and full regions (p = 0.04). No significant differences were observed for FAZ area, FAZ perimeter, or FAZ circularity (all p > 0.05).

**TABLE 2.**
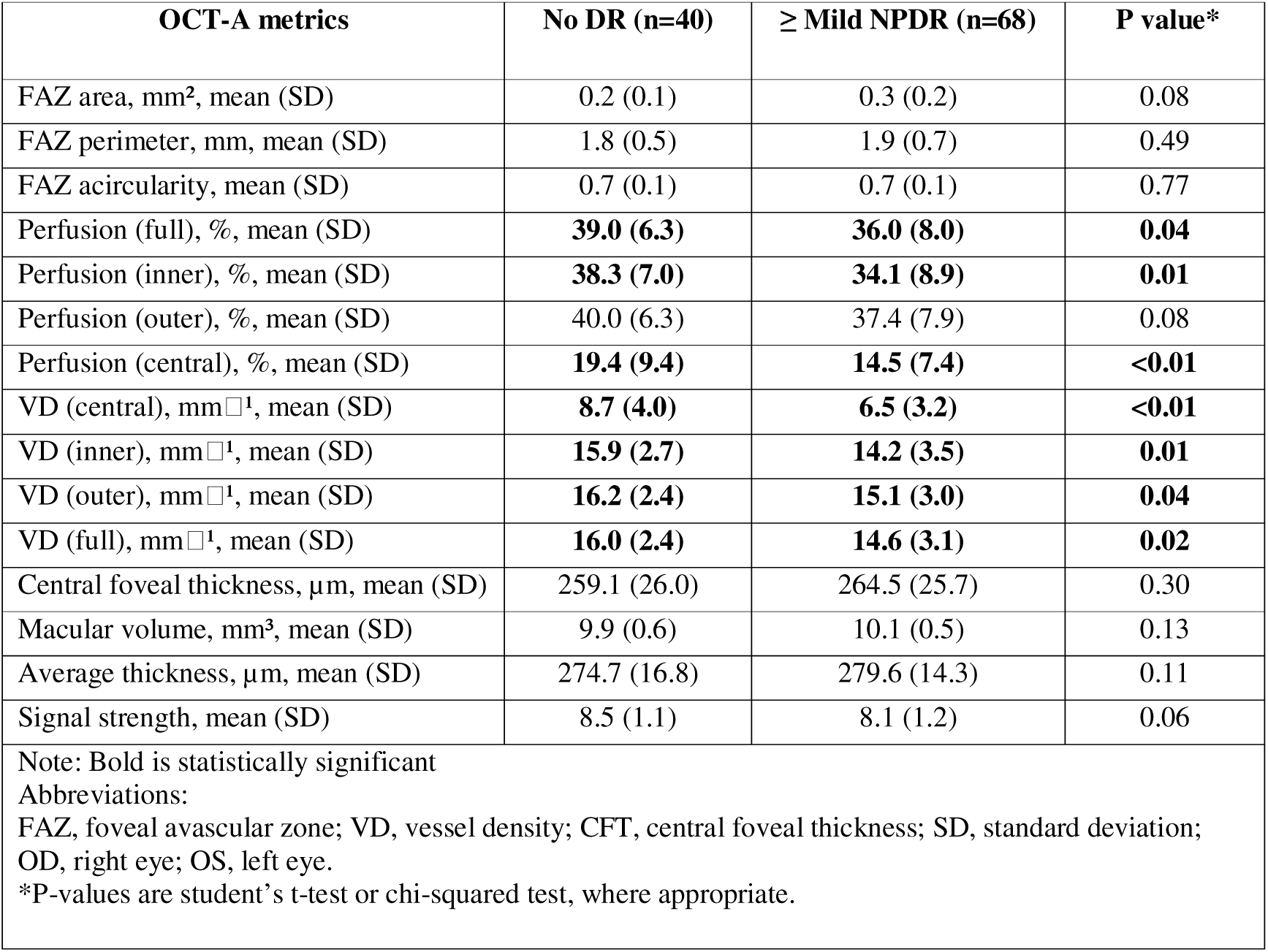
Optical coherence tomography angiography metrics in eyes without diabetic retinopathy and eyes with ≥ mild non-proliferative diabetic retinopathy.

### Multivariable logistic regression analysis for OCT-A parameters

After adjusting for age, sex, HbA1c, and diabetes duration, multivariable results (**Table 3**) showed lower VD in the central macula (OR = 0.82, 95% CI 0.68–0.98, p = 0.03) and reduced central PD (OR = 0.92, 95% CI 0.86–0.99, p = 0.03) remained independently associated with the presence of ≥ mild NPDR. Diabetes duration was also significantly associated with DR (OR = 1.05 per year, 95% CI 1.02–1.08, p < 0.001), while male sex was inversely associated (OR = 0.52, 95% CI 0.28–0.98, p = 0.04). We repeated analyses restricting to participants where the imaged eye was the worse eye (or equal); results were similar.

**TABLE 3:** Univariate and multivariable logistic regression analyses of optical coherence tomography angiography imaging biomarkers associated with the presence of diabetic retinopathy*.

| Variable | ≥ Mild NPDR OR<br>(95% CI)<br>Univariate | P value | ≥ Mild NPDR OR<br>(95% CI)<br>Multivariate* | P value |
| --- | --- | --- | --- | --- |
| **Demographic Factors** |  |  |  |  |
| Age (per decade) | 1.09 (1.08–1.11) | 0.26 | 1.09 (1.07–1.10) | 0.12 |
| Sex (Male) | **0.63 (0.39–1.02)** | **0.06** | **0.52 (0.28–0.98)** | **0.04** |
| **Clinical Factors** |  |  |  |  |
| HbA1c | 1.03 (0.94–1.14) | 0.52 | 1.04 (0.95–1.14) | 0.36 |
| Diabetes duration (years) | **1.03 (1.01–1.05)** | **0.01** | **1.05 (1.02–1.08)** | **<0.001** |
| **Imaging Biomarkers** |  |  |  |  |
| FAZ area (per 0.1mm<sup>2</sup>) | 1.01 (0.77–1.32) | 0.95 | 1.29 (0.78–2.16) | 0.32 |
| FAZ perimeter (mm) | 1.46 (0.75–2.83) | 0.26 | 1.59 (0.69–3.67) | 0.28 |
| FAZ acircularity | 0.43 (0.02–10.63) | 0.61 | 2.08 (0.04–100.68) | 0.71 |
| Perfusion full region (%) | 0.95 (0.89–1.00) | 0.06 | 0.95 (0.87–1.03) | 0.22 |
| Perfusion inner region (%) | **0.94 (0.88–0.99)** | **0.03** | 0.94 (0.87–1.02) | 0.15 |
| Perfusion outer region (%) | 0.95 (0.90–1.01) | 0.11 | 0.96 (0.88–1.04) | 0.28 |
| Perfusion central region (%) | **0.93 (0.88–0.98)** | **0.00** | **0.92 (0.86–0.99)** | **0.03** |
| Vessel density central region (mm<sup>-1</sup>) | **0.83 (0.74–0.94)** | **0.00** | **0.82 (0.68–0.98)** | **0.03** |
| Vessel density outer region (mm<sup>-1</sup>) | 0.86 (0.74–1.01) | 0.06 | 0.87 (0.70–1.08) | 0.21 |
| Vessel density full region (mm<sup>-1</sup>) | **0.84 (0.72–0.99)** | **0.04** | 0.85 (0.68–1.06) | 0.16 |
| Central foveal thickness, (μm) | 1.01 (1.00–1.02) | 0.20 | 1.01 (0.99–1.03) | 0.26 |
| Volume cube (mm<sup>3</sup>) | 1.90 (0.95–3.79) | 0.07 | 1.30 (0.51–3.29) | 0.58 |
| Average thickness (μm) | **1.02 (1.00–1.05)** | **0.06** | 1.01 (0.98–1.05) | 0.44 |
| Note: Bold is statistically significant<br>*Abbreviations:*<br>OR, odds ratio; CI, confidence interval; HbA1c, glycosylated haemoglobin; FAZ, foveal avascular zone; VD, vessel density; CFT, central foveal thickness; SD, standard deviation.<br>*Multivariable model adjusted for age, sex, HbA1c, and diabetes duration. |  |  |  |  |

### Logistic regression model performance

Logistic regression models evaluated the discriminative performance of systemic clinical variables and OCT-A parameters for the presence of ≥ mild NPDR (**Table 4 and Figure 2**). Model A used systemic clinical variables only (age, sex, HbA1c, and diabetes duration) and yielded an AUC of 0.60 (95% CI 0.45–0.75). In contrast, Model B solely used OCT-A parameters (central and inner VD and PD) and demonstrated superior discriminative ability with an AUC of 0.73 (95% CI 0.63–0.83). Model C combined clinical variables in Model A and OCT-A parameters in Model B and produced an AUC of 0.76 (95% CI 0.64–0.88).

**FIGURE 2:**
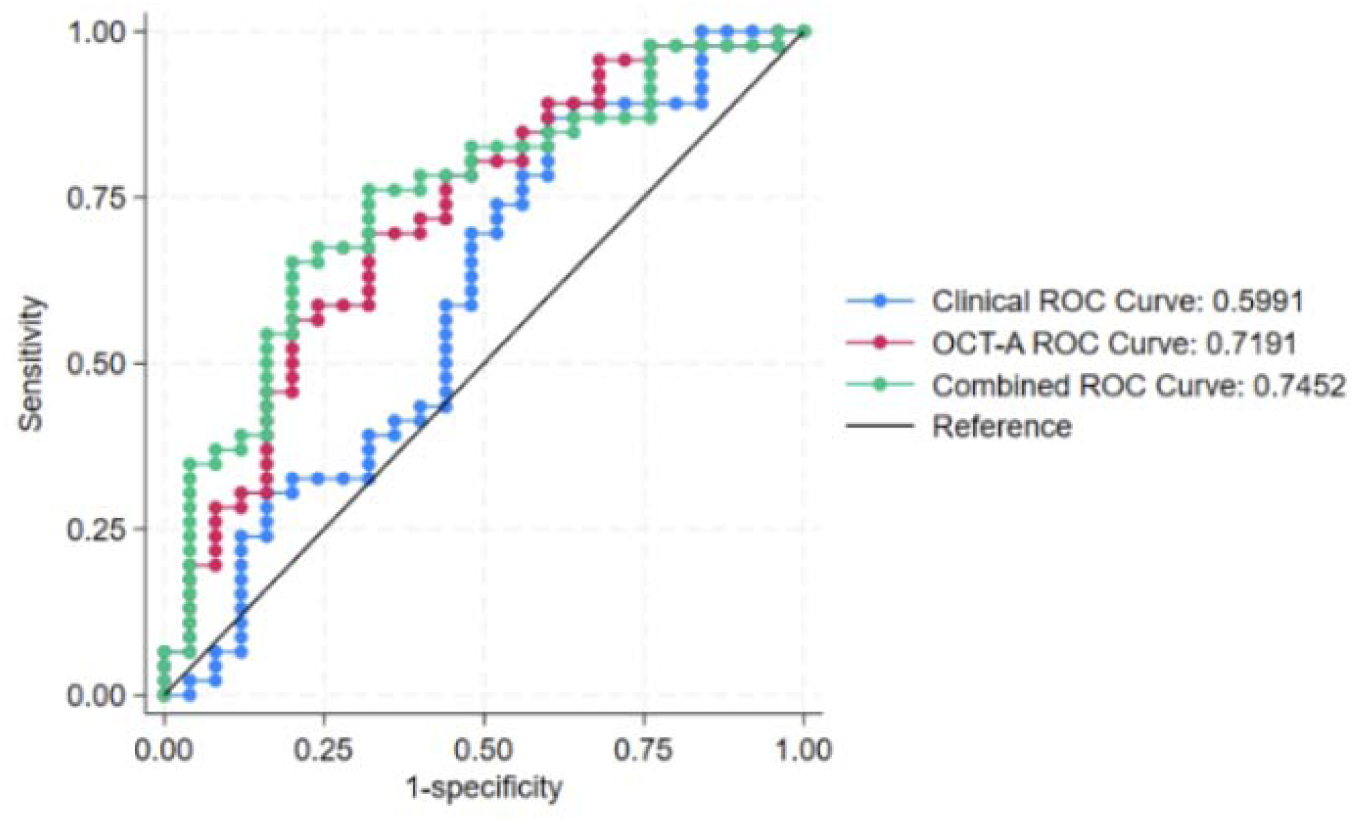
Receiver operating characteristic (ROC) curves for discriminating ≥ mild non-proliferative diabetic retinopathy using a clinical model (age, sex, diabetes duration, HbA1c), an OCT-A model (central and inner macular vessel and perfusion density), and a combined model.

**TABLE 4:** Logistic regression model performance for predicting the presence of ≥ mild non-proliferative diabetic retinopathy.

| Model | Predictors Included | AUC (95% CI) | Pseudo R<sup>2</sup> | Model $\chi^2$ (df) | P-value |
| --- | --- | --- | --- | --- | --- |
| **Model A** | Age, sex, HbA1c, diabetes Duration | 0.60 (0.45–0.75) | 0.06 | 16.85 (4) | **0.002** |
| **Model B** | VD central and inner, perfusion central and inner | 0.73 (0.63–0.83) | 0.12 | 17.30 (4) | **0.002** |
| **Model C** | Age, Sex, HbA1c, diabetes Duration + VD central and inner, perfusion central and inner | 0.76 (0.64–0.88) | 0.15 | 14.63 (8) | 0.067 |
| *Note: Bold is statistically significant*<br>*Abbreviations:*<br>*AUC, area under the curve; CI, confidence interval; R<sup>2</sup>, coefficient of determination; df, degrees of freedom; HbA1c, glycated haemoglobin; VD, vessel density.* |  |  |  |  |  |

## Discussion

In this cross-sectional observational study, 15 OCT-A parameters were examined in 108 eyes among participants with diabetes, including patients without DR and with ≥ mild NPDR. Eyes with ≥ mild NPDR showed significantly reduced VD in the central, inner, outer, and full macular regions, while PD was reduced in the central, inner, and full regions. FAZ parameters did not differ between groups, likely reflecting measurement variability, vendor-specific segmentation, and artefact-prone delineation. In the multivariable regression model, central VD and central PD were the strongest OCT-A predictors of DR presence. The ROC/AUC analysis showed OCT-A parameters alone provided greater discriminatory ability for detecting ≥ mild NPDR than routinely collected clinical factors, with the combined model offering only a modest incremental improvement.

In early DR, chronic hyperglycaemia activates multiple metabolic pathways, including the polyol pathway, which collectively heighten oxidative stress and pro-inflammatory signalling.^10^ These changes drive pericyte loss and endothelial cell damage, disrupting retinal neural-vascular interaction. This leads to basement membrane thickening, leukostasis, and accumulation of advanced glycation end-products, promoting retinal capillary dropout and non-perfusion.^10,11^ In this study, PD and VD in the SCP, reflecting retinal capillary health, were significantly reduced in eyes with DR compared to controls without DR, whereas retinal thickness and macular volume showed no significant differences. These findings suggest that impaired inner retinal circulation precede structural retinal change. The macula is particularly susceptible due to its high metabolic demand; thus, impaired perfusion can explain early retinal dysfunction seen in cases with no or minimal DR signs.^12^

The finding that OCT-A metrics (PD and VD) can detect DR presence is supported by Durbin et al.,^13^ who reported that the parafoveal VD acquired by the Zeiss AngioPlex OCT-A could reliably distinguish between eyes with no DR and early retinopathy. Similarly, several studies^14–18^ agree that OCT-A can detect subclinical microvascular compromise in patients with diabetes when compared to controls. However, like this study’s cohort, FAZ metrics as a stand-alone biomarker for detecting DR presence is less consistent within the literature,^18^ likely due to wide inter-individuality variability and segmentation challenges. FAZ variability reinforces the study’s results that VD and PD are stronger markers for DR detection. Much of the existing OCT-A literature^17,19^ has emphasised DR severity or progression rather than binary detection. While these studies are clinically relevant in monitoring DR stages, differentiating between the presence and absence of DR is yet to be explored. This distinction underscores a gap in the literature, as OCT-A is validated for grading DR severity, yet few studies have evaluated its diagnostic utility for detecting DR.

Early risk stratification in DR is essential for timely interventions. Traditional systemic risk factors, including age, sex, HbA1c, and duration of diabetes, are associated with higher DR risk;^20^ higher HbA1c and longer diabetes duration has consistently predicted DR development.^21,22^ OCT-A metrics show strong utility for grading DR severity and monitoring progression in the SCP and DCP.^11,13,22–25^ However, few studies have directly compared the discriminative ability of these systemic factors with OCT-A metrics. In this study, SCP metrics, particularly central and inner VD and PD, demonstrated superior discriminative performance in detecting DR presence compared to systemic variables (Fig. 2.). Therefore, this study suggests that OCT-A metric alone have the potential to discriminate the presence of DR, with potential to complement, or in some settings, replace conventional systemic indicators.

The clinical relevance of these findings lies in the potential application of OCT-A metrics as adjunctive tools for the early detection of DR and associated risk factors. The ability of a 6×6 mm OCT-A scan to visualise and quantify microvascular changes provides an opportunity to shift detection thresholds to earlier disease stages, with VD and PD parameters showing clinically meaningful changes within this central zone, even though early DR alterations are more clinically observed in the periphery.^26^ Our findings demonstrate that macular metrics can still provide discriminative value for DR detection in the absence of CSMO, highlighting the utility of OCT-A for identifying microvascular compromise without the need to go beyond the 6×6 mm scan. The study’s results also show that a larger FAZ metric can contribute to a composite risk profile, when considered alongside reductions in VD and PD, rather than serving as an isolated diagnostic marker.^27^ These OCT-A metrics are therefore of clinical importance, as earlier detection of diabetic microvascular compromise offers the potential for closer monitoring, systemic management and earlier ophthalmic intervention. The non-invasive nature of OCT-A also enhances its feasibility for longitudinal monitoring.

This study’s major strength lies in its use of quantitative OCT-A metrics to detect early microvascular changes in DR, enabling binary identification of disease presence at early stages. Additionally, the study includes rigorous image-quality controls such as image quality thresholds (signal strength >7), tracker-assisted acquisition, and predefined exclusions for motion, projection, and segmentation artefacts, which minimised measurement bias and improved the reliability of the OCT-A metrics.

Our study also has several limitations. Firstly, the analysis of OCT-A metrics was restricted to the SCP, and the DCP was not captured. Secondly, although patients with CSMO were excluded, 14.7% of the cohort with ≥ mild NPDR had non-clinically significant macular oedema, which may have influenced vascular and thickness metrics. Thirdly, subgroup comparisons between patients with type 1 and type 2 diabetes was constrained, limiting further statistical analysis and generalisability of the findings. The FAZ parameters showed no independent association with DR, which may reflect measurement variability or inter-correlation with other OCT-A metrics. Additionally, as the analysis used a participant-level outcome but single-eye OCT-A imaging, some degree of misclassification between eyes cannot be ruled out. Furthermore, as the AngioPlex segmentation algorithms and signal threshold are vendor-specific, generalisability to other OCT-A platforms may be limited. Finally, signal strength was slightly lower in DR group, however, it was above the quality threshold in all scans and unlikely to have affected the results.

As a cross-sectional analysis, this study cannot determine temporal ordering or progression; associations between OCT-A metrics and DR presence should not be interpreted as causal, nor as predictors of future change. Therefore, larger, longitudinal studies capturing both SCP and DCP across a broader range of disease severity are required to validate OCT-A metrics as robust biomarkers of DR disease progression, and future work should include cross-platform validation to confirm parameter comparability.

## Conclusion

This study demonstrated that OCT-A-derived VD and PD in the SCP are independently associated with the presence of early DR. These OCT-A metrics significantly outperformed traditional clinical risk factors in distinguishing eyes with ≥mild NPDR, from those with no or minimal DR, highlighting the potential for OCT-A metrics in facilitating the clinical decision-making and early detection. Further longitudinal studies are warranted to validate these findings across broader populations and stages of disease progression.

## Data Availability

The data that support the findings of this study are not publicly available due to patient privacy and ethical restrictions but are available from the corresponding author upon reasonable request, subject to approval by the relevant ethics committee.

## Acknowledgements

The authors thank the clinical teams at the Flinders Optometry and Ophthalmology Clinics for their assistance with patient recruitment and data collection.

## Declaration of interest statement

The authors report there are no competing interests to declare.

## Journal type

Original research paper

## Journal submission

Ophthalmic Epidemiology

## Submission status

This manuscript is original, not under consideration or published elsewhere, and has been approved by all authors.

## Funding

This research received no specific grant from any funding agency in the public, commercial, or not-for-profit sectors.

## Author contributions

JE, KK, MH and AH were responsible for the study design. Data analysis was performed by JE, EP and AH. All authors contributed to drafting the manuscript, interpreting the findings, critically revising the content, and approving the final version for submission.

## Conflict of interests

The authors declare no conflicts of interest.

## Notes

### Competing Interest Statement

The authors have declared no competing interest.

### Author Declarations

Ethical approval was obtained from the Southern Adelaide Local Health Network Human Research Ethics Committee (#HRE00228). All participants received written and verbal information about the study and provided written consent. The study adhered with the National Statement on Ethical Conduct in Human Research and the Declaration of Helsinki.

